# Summary statistics from large-scale gene-environment interaction studies for re-analysis and meta-analysis

**DOI:** 10.1101/2023.05.08.23289686

**Authors:** Duy T. Pham, Kenneth E. Westerman, Cong Pan, Ling Chen, Shylaja Srinivasan, Elvira Isganaitis, Mary Ellen Vajravelu, Fida Bacha, Steve Chernausek, Rose Gubitosi- Klug, Jasmin Divers, Catherine Pihoker, Santica M. Marcovina, Alisa K. Manning, Han Chen

## Abstract

Summary statistics from genome-wide association studies enable many valuable downstream analyses that are more efficient than individual-level data analysis while also reducing privacy concerns. As growing sample sizes enable better-powered analysis of gene-environment interactions (GEIs), there is a need for GEI-specific methods that manipulate and use summary statistics. We introduce two tools to facilitate such analysis, with a focus on statistical models containing multiple gene-exposure and/or gene-covariate interaction terms. REGEM (RE-analysis of GEM summary statistics) uses summary statistics from a single, multi-exposure genome-wide interaction study (GWIS) to derive analogous sets of summary statistics with arbitrary sets of exposures and interaction covariate adjustments. METAGEM (META-analysis of GEM summary statistics) extends current fixed-effects meta-analysis models to incorporate multiple exposures from multiple studies. We demonstrate the value and efficiency of these tools by exploring alternative methods of accounting for ancestry-related population stratification in GWIS in the UK Biobank as well as by conducting a multi-exposure GWIS meta-analysis in cohorts from the diabetes-focused ProDiGY consortium. These programs help to maximize the value of summary statistics from diverse and complex GEI studies.

## Introduction

Gene-environment interaction (GEI) analysis is a key tool for understanding genetic impacts on human traits, with the potential to account for additional heritability, explain differences in genetic effects across populations, and support personalized lifestyle and therapeutic decisions. Historically, GEI studies have taken a hypothesis-driven approach, but larger cohorts,^1^ and new software programs have provided the necessary statistical power and computational efficiency to study GEIs genome-wide.^2–7^ These genome-wide interaction studies (GWIS) generate summary statistics, or variant-level regression results, which have substantial value beyond locus mapping. For example, summary statistics allow for heritability analysis,^8^ enrichment testing,^1^ and genome-wide polygenic score generation.^1, 9^

GEI analysis and interpretation are complicated by the densely correlated set of possible exposures that may interact with genotypes to influence human traits (the “exposome”, defined here as including demographic and physiologic traits). Two modeling implications are particularly pertinent. First, multi-exposure GEI analysis can increase statistical power by jointly testing genetic interactions with multiple exposures.^5, 10, 11^ This strategy can pool signals across distinct exposures (e.g., smoking status and pollution exposure for lung function) or incorporate multiple definitions of a single exposure category (e.g., current smoking status and pack-years of smoking). Second, proper control of confounding for GEI interaction terms requires adjustment for not just the main effects of covariates, but also their genetic interactions.^12^ Inclusion of these “interaction covariates” is thus necessary to produce interpretable summary statistics.

Rigorous GEI analysis carries complexities stemming from its place at the center of traditional and genetic epidemiology. Sensitivity analyses, while commonplace in traditional epidemiology, are computationally burdensome when conducted across millions of variants genome-wide. Meanwhile, well-established meta-analysis procedures for genome-wide association study (GWAS) summary statistics become more difficult in the context of multi-exposure GEI models. Software programs do not yet exist to perform efficient meta-analysis in the context of these complex analytical designs.

We introduce methods and associated software programs to advance the field of genome-wide GEI analysis based on summary statistics. While the statistical results are general, the associated software implementations build on the results from our previously described software program for efficient GWIS, GEM.^2^ Exploiting the redundancy of statistical estimates across related GEI models, we introduce the REGEM (RE-analysis of GEM summary statistics) program to derive genome-wide summary statistics corresponding to arbitrary multi-exposure and interaction covariate adjustments based on results from a single, multi-exposure GWIS. Expanding current fixed-effect meta-analysis models, we further introduce the METAGEM (META-analysis of GEM summary statistics) program to conduct efficient meta-analysis of GEI effects under complex GEI analysis models. We demonstrate the value and efficiency of these tools by exploring alternative methods of accounting for population stratification in GWIS in the UK Biobank as well as by conducting a multi-exposure GWIS meta-analysis in cohorts from the ProDiGY consortium.

## Material and methods

### GEM method

We developed two C++ software programs that use summary statistics from GEI studies. REGEM requires output from a single GEI study, while METAGEM requires output from multiple GEI studies. Both programs are designed for easy integration with output from GEM. Here we summarize the GEM methodology. For a single-variant test of N unrelated individuals, GEM considers the generalized linear model:

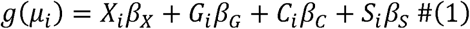

for individual *i*, where 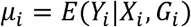 is the conditional mean of the phenotype *Y_i_* given *p* covariates *X_i_* (including the intercept), and the genotype *G_i_* for a single genetic variant. The interaction terms *C_i_* and *S_i_* are the products of *G_i_* and *c* covariates and *q* exposures (which are disjoint subsets of *X_i_*), respectively.^2^ Let *Y* = (*Y*_1_ *Y*_2_ ... *Y_N_*)^*T*^ be a length *N* vector of phenotypes, 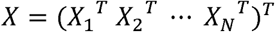 be an *N* x *p* matrix of *p* covariates, *G* = (*G*_1_ *G*_2_ ... *G_N_*)^*T*^ be a length *N* vector of genotypes for this single genetic variant, 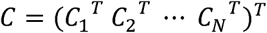 be an *N* x *c* matrix of *c* gene-covariate interaction terms, 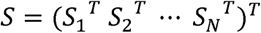 be an *N* x *q* matrix of *q* gene-environment (exposure) interaction terms, we can fit a null model without any genetic effects *g*(*Ε_i_*) = *X_i_β_x_* and get a length *N* residual vector *r*. Let 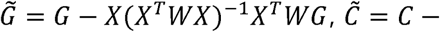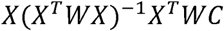 and 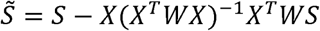 be covariate *X* adjusted *G, C* and *S*, respectively, where *W* is a diagonal weight matrix with elements 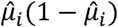 for logistic regressions (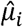 are fitted probabilities of *Y_i_* = 1 from the null model) and an identity matrix for linear regressions, GEM computes a length (1 + *c* + *q*) score vector 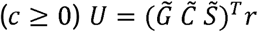, and (1 + *c* + *q*) x (1 + *c* + *q*) matrices 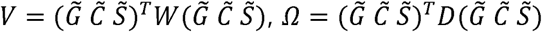, where *D* is a diagonal matrix of squared residuals.

For *M* variants in a genome-wide scan, we retrieve the dispersion parameter estimate, 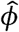(which is fixed at 1 for logistic regressions and the residual variance estimate from the null model for linear regressions), the genetic main effect, gene-covariate interaction effects and gene-environment (exposure) interaction effects, as well as both model-based and robust standard errors and covariances for *G*, *C* and *S*. The effect estimates are computed as 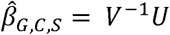. The full (1 + *c* + *q*) x (1 + *c* + *q*) model-based and robust variance-covariance matrices are computed as 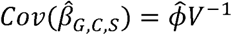 and 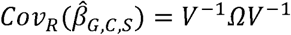, respectively. In the full output, GEM (version 1.3 and later) reports the model-based and robust standard errors of effect estimates, which are the square root of the diagonal elements of 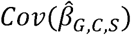 and 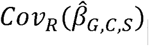, as well as the model-based and robust covariances for these effect estimates (the off-diagonal elements of 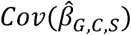 and 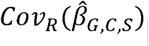.

### REGEM Method

Given the full summary statistics output from GEM (version 1.3 and later), the score vector and matrices *V* and *Ω*, can be reconstructed without access to individual-level data. Utilizing 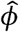 and the matrices 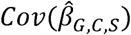 and 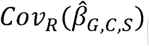 described above, it follows that 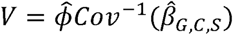 and 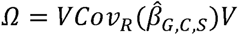. The score vector can then be recomputed as 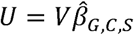.

REGEM supports two scenarios for re-analysis of a single GEI study. The first scenario involves the exclusion of one or more gene-covariate or gene-environment interaction terms from the original model. This is achieved by filtering *U* to exclude the specified gene-covariate or gene-environment interaction terms, resulting in the modified score vector 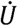. Subsequently, the matrices *V* and *Ω* are reduced to exclude the corresponding rows and columns of the specified gene-covariate or gene-environment interaction terms, denoted 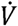 and 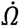. The GEM method can then be applied to 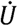, 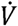, and 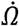 to obtain new summary statistics. In the second scenario, re-analysis can be performed by conditioning on one or more gene-environment interaction terms in the original GEM analysis as gene-covariate interactions or testing one or more gene-covariate interaction terms in the original GEM analysis as gene-environment interaction terms of interests. In either case, the ordering of *U* is rearranged, denoted as 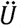 , to incorporate the original gene-environment interaction terms into *C* or the original gene-covariate interaction terms into *S*. The rows and columns of the matrices *V* and *Ω* are also reordered and denoted as 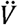 and 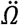. The GEM method follows for 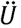, 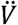, and 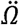. Both scenarios can be applied simultaneously.

### METAGEM method

METAGEM combines summary statistics from *K* independent studies using the inverse-variance weighted approach. For individual studies *k* = 1, 2, .. , *K*, with effect estimates 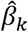 and the variance-covariance matrix *Cov_k_* from the GEM output (model-based or robust), the summary effect estimates are computed as 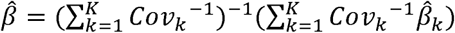, with the model-based or robust variance-covariance matrix 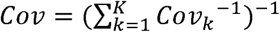.

### REGEM Comparison and Benchmark

To demonstrate the computational benefits of REGEM, we test and compare four variations of the waist-hip ratio (WHR) model originally described by Westerman et al. The original model is defined as follows (excluding the array covariate and PC6 - PC10):

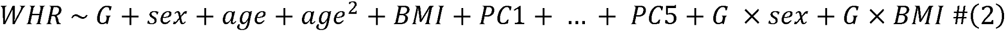

where WHR is the phenotype, sex is the primary exposure of interest, BMI is the interaction covariate, and age, age^2^, and PC1-PC5 are the covariates. Here, we retrieved PCs calculated as part of the Pan-UKBB project.^13^ All terms in the model were centered. First, we performed a genome-wide analysis of the original model using GEM (version 1.5) using 362,449 unrelated European-ancestry participants, and filtered variants with minor allele frequency (MAF) < 0.001, leaving 16,539,280 variants for re-analysis. Next, we derived associated genome-wide summary statistics corresponding to variations of the original model using REGEM, comparing their results and runtimes to simply re-running that same model genome-wide using GEM. Table S1 summarizes the variations of the original models, including the original model. These variations involve the joint testing of G x sex and G x BMI (M1), testing for G x BMI while adjusting for G x sex (M2), testing for G x sex while removing the G x BMI term (M3), and testing for G x BMI while removing the G x sex term (M4). All analyses were performed on the DNAnexus platform using the *mem1_ssd1_v2_x16* instance type, and we reported the runtime and memory usage of each run. The GEM and REGEM summary statistic comparisons were visualized using the scattermore and ggplot2 R packages.

### METAGEM Comparison and Benchmark

To evaluate the computational efficiency of METAGEM, we conducted a simulation study using phenotype and genotype data from the Pan-UKBB.^13^ We randomly split the phenotype data, which comprised 362,449 samples, into 11 datasets: one with 100,000 samples, two with 50,000 samples, seven with 10,000 samples, and one with 92,449 samples. For each dataset, we conducted a genome-wide gene-sex interaction test and filtered out variants with a MAF < 0.001, resulting in 15.46 to 16.85 million variants per dataset, and a total of 17,993,341 unique variants across all datasets. We then performed a gene-sex interaction meta-analysis using METAGEM and the METAL software (version 2010-02-08),^14^ with the joint meta-analysis patch,^15^ and compared the results. Additionally, we conducted a genome-wide joint gene-sex and gene-BMI interaction test for each dataset and performed a meta-analysis using METAGEM to evaluate its performance in the presence of multiple interaction terms. All analyses were conducted on the DNAnexus platform using a single core and the *mem1_ssd1_v2_x16* instance type. We reported the CPU time and memory usage for each analysis. We used the scattermore and ggplot2 R packages to visualize the comparison of summary statistics between METAGEM and METAL.

### Multi-exposure interactions influencing waist-hip ratio in the UK Biobank

Expanding the WHR analyses described above, we performed multiple GWIS, with downstream analysis using REGEM and METAGEM, to investigate genetic interactions with sex and BMI across multiple ancestries. The primary model, run using GEM, was conducted in unrelated individuals from multiple ancestries (N = 379,092) and followed model (2) above with the addition of gene-ancestry interaction covariates. Ancestry labels (AFR, AMR, CSA, EAS, EUR, and MID) were retrieved from the Pan-UKBB effort and were coded using five indicator variables, with EUR as the reference group. Using REGEM, we then derived summary statistics corresponding to equivalent single-exposure GWIS in the pooled-ancestry sample (testing only gene-sex or only gene-BMI interactions, while adjusting for only the main effect of the other). Additionally, we ran ancestry-stratified, multi-exposure analyses (using the same model but removing all covariate and interaction covariate terms containing ancestry labels). These ancestry-stratified analyses were then combined using METAGEM to generate meta-analyzed, multi-exposure interaction tests for comparison to the results from the ancestry-pooled analysis.

To compare locus discoveries across analysis strategies (e.g., ancestry-pooled vs. cross-ancestry meta-analysis), we first independently clumped summary statistics from each analysis using a distance-based method that grouped variants within 500kb of each lead variant. We then concatenated the clumped results from the two analyses and performed a secondary clumping using the same strategy, such that clumped loci in this second stage were considered to represent the same locus.

### Progress in Diabetes Genetics in Youth (ProDiGY) dataset

ProDiGY is a multi-ethnic resource including three studies: Treatment Options for Type 2 Diabetes in Adolescents and Youth (TODAY),^16^ SEARCH for Diabetes in Youth (SEARCH),^17^ and T2D-GENES. In total, the dataset contains 2,820 youth and 4,858 adult cases with T2D, and 656 diabetes-free youth and 4,934 adult controls after removing individuals with maturity-onset diabetes of the young (MODY) and type I diabetes. Samples were genotyped on the Infinium GWAS array by the Genetic Analysis Platform at the Broad Institute of MIT and Harvard. Details on quality control procedures for the genotype data have been previously described.^18^ Genotype data were imputed on the TOPMed Imputation Server using the TOPMed v2 reference panel. Variants passing an imputation quality threshold (R^2^) of 0.5 were retained for analysis. Genetic ancestry groups were assigned to ProDiGY samples based on genetic principal components analysis after merging with the 1000 Genomes dataset.

### Application multi-interaction to T2D in ProDiGY

To show the performance of METAGEM in the multi-gene-environment interactions with a real and genome-wide study, we first used GEM to conduct a multi-exposure gene-sex and gene-age interaction analysis for incident T2D, separately within each genetic ancestry group in two different comparisons: youth cases vs. youth controls (youth group) and adult cases vs. adult controls (adult group). Sex and age were both used as exposures and tested jointly for interaction using robust standard errors. Covariates included age, sex and 10 genetic principal components.

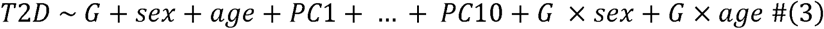

Using the full output from GEM, we performed cross-ancestry meta-analysis using METAGEM in both youth group and adult group analyses. We also conducted equivalent single-exposure GWIS with sex and age separately for comparison with the multi-exposure scan. Meta-analysis for these single-exposure tests was conducted using METAL, for both the joint (genetic plus interaction effect) test (patched version 2010-02-08; the only version for which the patch is available) and marginal test (version 2011-03-25) to conduct the marginal meta-analysis test across genetic ancestry groups. A threshold of *p*<5 ×10^−8^ was used to define genome-wide significance.

## RESULTS

Figure 1 shows the suite of software tools described here in the context of an analysis workflow, along with an example set of associated statistical models.

**Figure 1:**
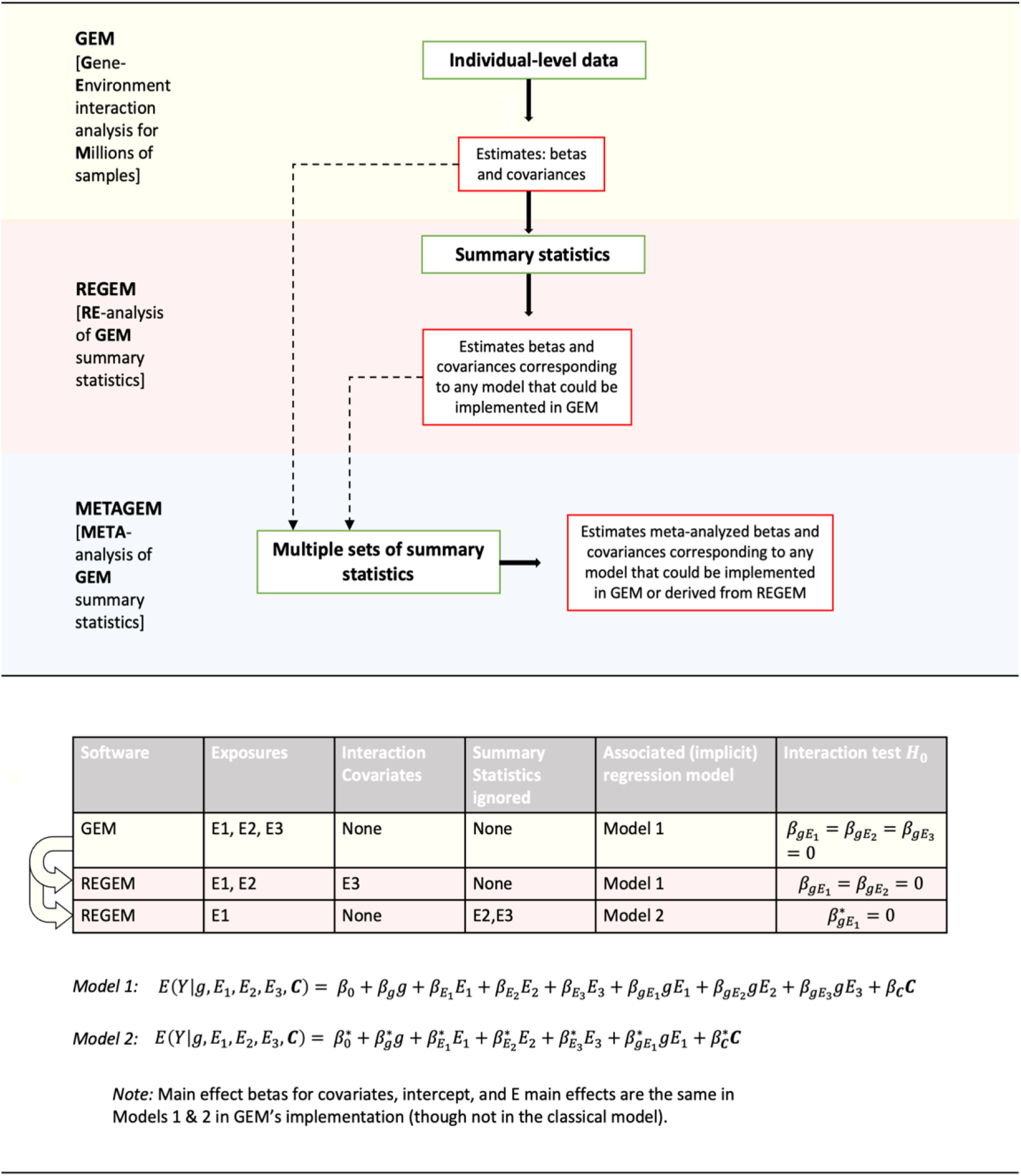
Large-scale GxE methods software suite and connections in the context of an analysis workflow. GEM (previously published) conducts genome-wide interaction studies for single datasets. Given multi-exposure summary statistics from GEM (version 1.3 and later), REGEM can estimate genome-wide summary statistics from an associated model that re-partitions any subset of exposures into interaction covariates and simple main effect adjustments without interaction. Given multiple sets of summary statistics from GEM and/or REGEM, METAGEM conducts meta-analysis for any number of jointly-tested exposures and interaction covariates.

### REGEM computational performance

We compared results obtained from genome-wide interactions tests using the REGEM and GEM methods across four distinct GEI models. The benchmark results, presented in Table 1, indicate that REGEM significantly reduces CPU time by eliminating the need for computation on individual-level data. For each model, REGEM completed a genome-wide run in less than 6 minutes, while GEM required several CPU days to achieve the same outcome. Additionally, re-analyses for multiple interactions (M1 and M2) using REGEM took only about a minute of additional CPU time compared to single exposure re-analyses (M3 and M4). Overall, REGEM saved considerable time, ranging from hours to days of computation time. Moreover, the memory requirements for REGEM were minimal, primarily depending on the number of gene-environment interaction terms, which are usually small. Finally, the effect and variance estimates from REGEM were consistent with those obtained from GEM for each of the four models (M1-M4) as shown in Figures S1-S4.

**Table 1.** Genome-wide re-analysis benchmark comparison between GEM and REGEM.

| Benchmark | GEM |  |  |  | REGEM |  |  |  |
| --- | --- | --- | --- | --- | --- | --- | --- | --- |
|  | M1 | M2 | M3 | M4 | M1 | M2 | M3 | M4 |
| CPU time (Mins) | 13,972.17 | 13,618.44 | 10,959.33 | 10,994.26 | 5.22 | 5.20 | 4.43 | 4.06 |
| Memory (MB) | 2,325.37 | 2,342.48 | 2,188.14 | 2,188.14 | 13.66 | 13.64 | 11.43 | 11.63 |

### METAGEM computational performance

Genome-wide meta-analysis runs of ∼17.99 million variants, derived from 11 simulated UKB datasets, were carried out using the METAGEM and METAL methods with a single core. Table 2 summarizes the CPU time and memory usage of the runs. For a single exposure meta-analysis, METAGEM showed a modest improvement in performance compared to METAL, completing the run approximately 2 minutes faster and using approximately 1 GB less memory. We note that METAGEM meta-analyzed all 17,993,341 variants, while METAL skipped 25,670 multi-allelic variants that contained duplicate variant identifiers. However, the impact of the skipped variants on the benchmark results was negligible. Model-based and robust meta-analysis results from METAGEM and METAL are compared in Figure S5. As expected, the summary statistics and joint *P*-values were consistent between the two methods. To test the performance of METAGEM in conducting meta-analysis with multiple interactions, we performed genome-wide joint meta-analysis with gene-sex and gene-BMI as the interactions using METAGEM. As shown in Table 2, METAGEM efficiently completed the run in an additional ∼6 minutes of CPU time and less than 1 GB of additional memory compared to the single exposure meta-analysis.

**Table 2.**
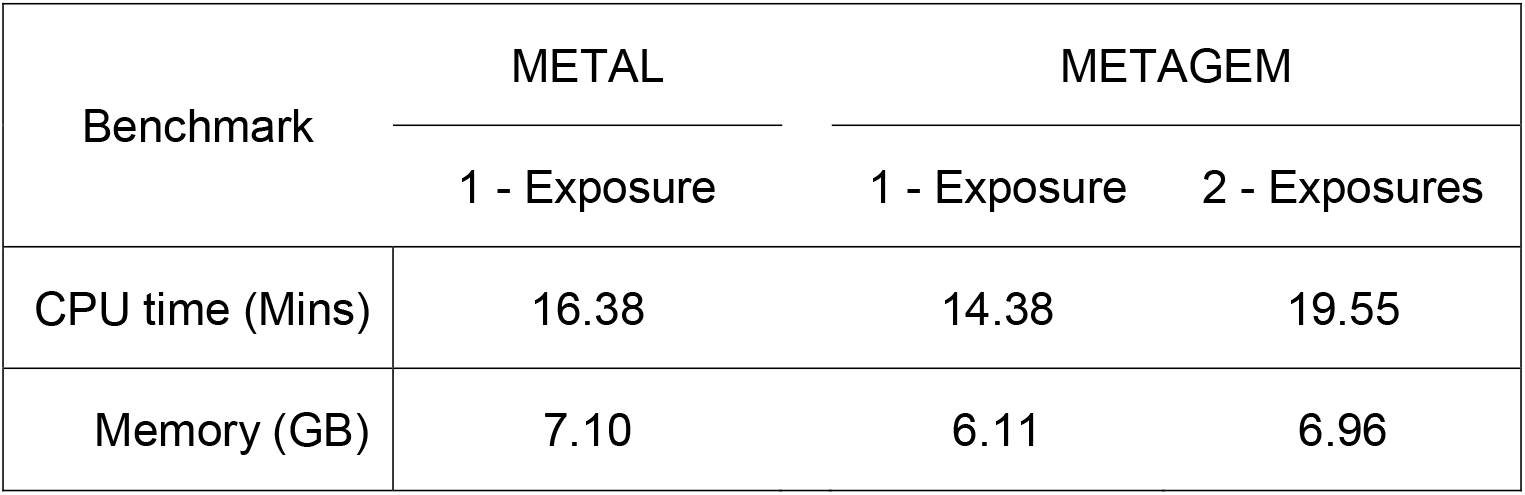
Genome-wide meta-analysis benchmark between METAL and METAGEM for 17,993,341 variants using a single core.

### Accounting for ancestry in pooled analysis of waist-hip ratio

In order to test the functionality of REGEM and METAGEM on real datasets, we further explored the expanded WHR GWIS model used for benchmarking. The primary analysis tested genetic interactions with two exposures (sex and BMI) in a pooled dataset containing six ancestry groups. Without additional adjustments, this pooled dataset produced highly inflated summary statistics (genomic inflation lambda = 5.35), but after inclusion of interaction covariates (gene-ancestry and exposure-ancestry interaction terms), this inflation was reduced to a level identical to that of a European ancestry-only analysis (lambda = 1.18 for both; Figure 2a). This properly-adjusted pooled analysis uncovered 55 independent loci using a standard genome-wide significance threshold of 5×10^−8^. Using REGEM to produce equivalent single-exposure interaction tests (sex or BMI), we saw that the sex-only GWIS revealed a highly overlapping set of loci (57 loci in total, 47 of which overlapped loci from the multi-exposure test), while the BMI-only GWIS revealed many fewer (6 loci in total, 5 of which overlapped loci from the multi-exposure test; Figure 2b).

**Figure 2:**
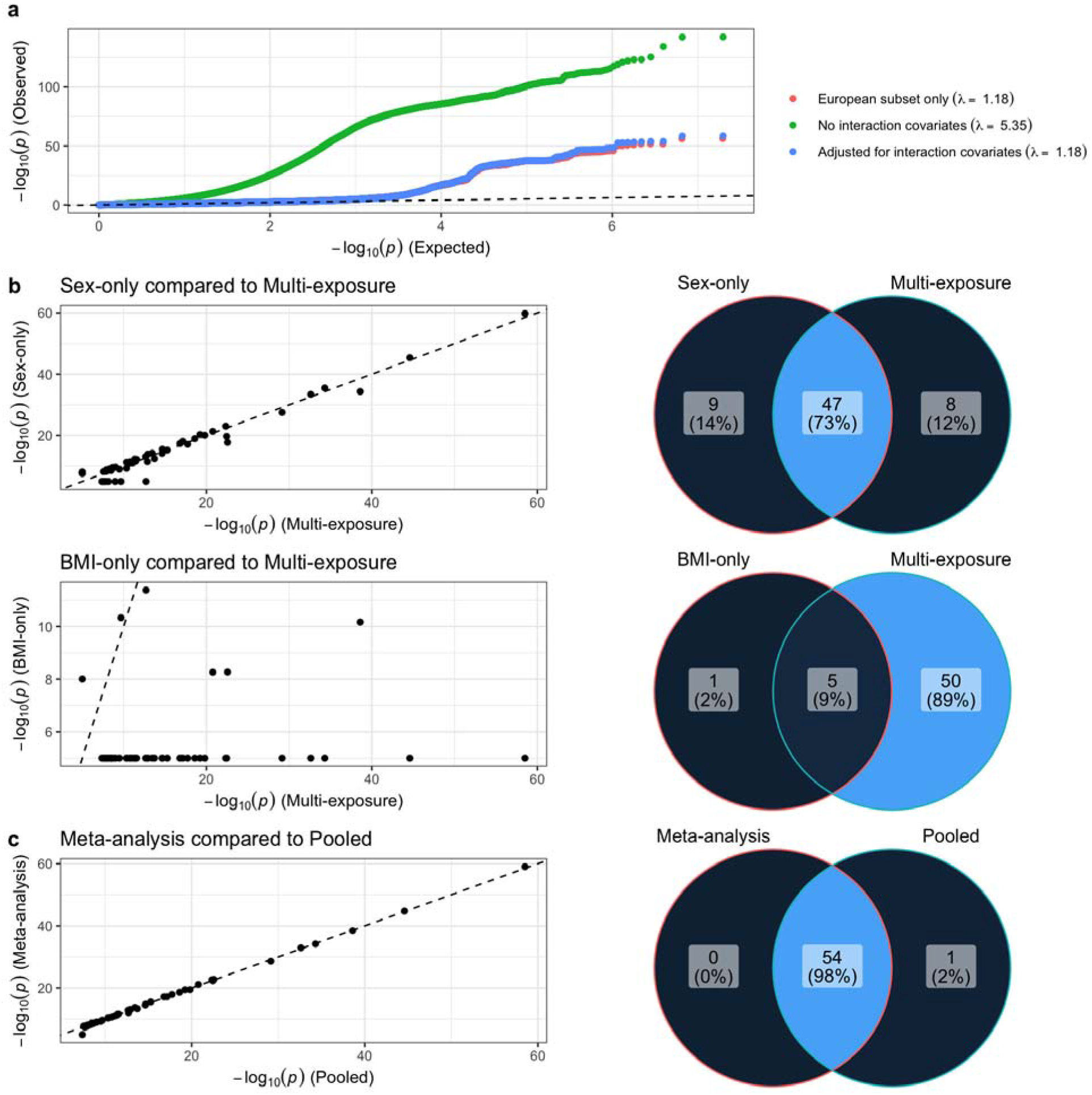
Results from multi-exposure, multi-ancestry GWIS for waist-hip ratio. a) Quantile-Quantile plots display observed vs. expected p-values for selected analyses. b) Results from REGEM-derived, single-exposure GWIS results for sex (top panel) and BMI (bottom panel). Scatter plots compare p-values between single- and multi-exposure interaction tests and Venn diagrams display the overlap in independent loci discovered using single- and multi-exposure interaction tests. c) As in (b), but replacing REGEM-derived, single-exposure results with METAGEM-derived, multi-ancestry meta-analysis results.

Using METAGEM, we then conducted a meta-analysis of six ancestry-specific GWIS, finding 54 total loci, all of which overlapped loci from the primary ancestry-pooled analysis (Figure 2c). This high concordance reinforces two conclusions. First, proper adjustment for interaction covariates can allow rigorous pooled-ancestry GWIS and avoid the need for stratification. Second, in situations where pooled analysis is not possible for logistical or analytical reasons, the ability to adjust for interaction covariates and possibly include multiple exposures in conducting GWIS meta-analysis can be critical for proper interpretation and control of inflation.

### Sex and age interaction effects on T2D in the ProDiGY dataset

We performed a genome-wide, multi-exposure test of sex and age interactions affecting T2D analysis in the ProDiGY dataset, separately in the youth (youth cases vs youth controls) and adult (adult cases vs. adult controls) subsets. After cross-ancestry meta-analysis, we did not detect any significant signals using the interaction test, but using the joint test found 8 independent loci passed the genome-wide significance threshold in the youth group (Table S2) and 3 loci in the adult group (Table S3). Of the 8 loci in the youth group, two were known associations, at *TCF7L2* (*p_joint_* = 1.30×10^−9^) and *MC4R* (*p_joint_* = 9.22×10^−9^). Only one, rs7903146 at *TCF7L2*, showed a significant effect in the marginal genetic effect test (excluding interaction effects). Six of the 8 signals were not reported in previous T2D GWAS studies (as per the Common Metabolic Disease Knowledge Portal). One variant, rs114578532, upstream of *FGF6*, passed the genome-wide significance threshold in the marginal test (*p_marginal_*= 2.18×10^−8^), but not joint test (*p_joint_*= 7.25×10^−7^). These signals, with the exception of *TCF7L2*, did not show strong effects in the adult group analysis. In the adult cases vs. adult controls comparison, out of three signals, two were known to be associated with T2D and also showed statistical significance in the marginal test (rs35198068 at *TCF7L2* and rs2237892 at *KCNQ1*). The third locus, with lead variant rs62287662 within an intron of *KCNAB1*, has not been previously associated with T2D (*p_joint_*= 1.79×10^−8^; *p_interaction_*= 6.27×10^−8^). *KCNAB1* encodes a protein involved in diverse functions including heart rate and insulin secretion. This locus did not show meaningful association in the youth group analysis.

To evaluate the added value of multi-exposure analysis, we ran analogous single-exposure meta-analyses, separately for sex and age. Of 8 multi-exposure signals in the youth group joint test, we found that 5 reached significance in the sex-only analysis (plus 2 additional signals) and 3 in the age-only analysis (plus 1 additional signal) (Figure 3). In the adult group, 2 of 3 loci were found in all three models, with the third found in both the multi-exposure and age-only tests but not the sex-only test (Figure S6).

**Figure 3:**
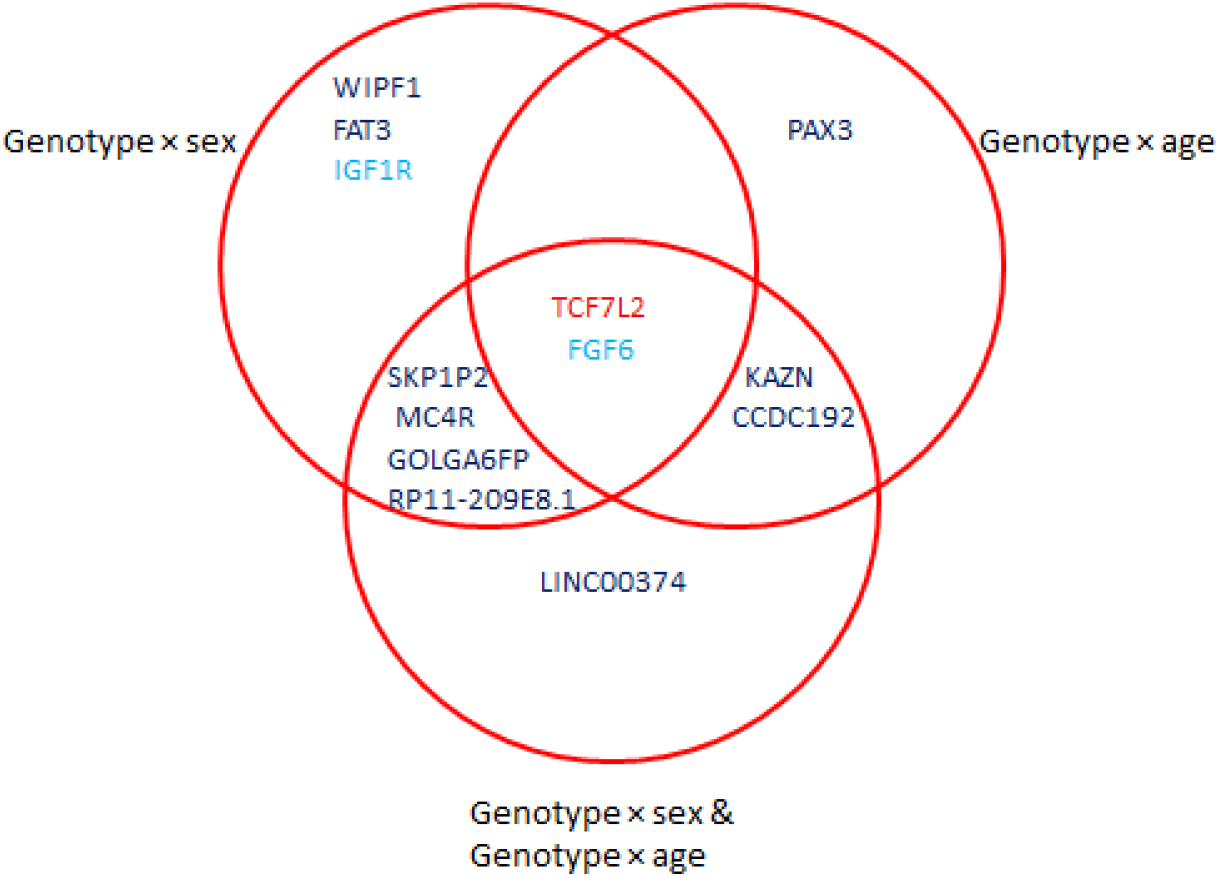
Results from multi-exposure GWIS for incident T2D in the ProDiGY youth cohort. Venn diagram displays overlap between loci discovered at genome-wide significance using the joint test of genetic and interaction effects (*p_joint_* = 5×10^−8^), from each of: sex-only, age-only, and multi-exposure (sex and age) analyses. Variants are labeled according to the closest gene, and colors correspond to the test(s) in which significance was achieved: marginal genetic effect (light blue), joint genetic effect (dark blue), or both joint and marginal genetic effects (red).

## DISCUSSION

GEI studies are becoming increasingly challenging due to complex structured models involving multiple interaction terms. Here we introduce two software programs, REGEM and METAGEM, to enable further downstream analysis of such studies using only summary statistics. We show that both programs are much more computationally efficient than the corresponding individual-level data analyses and validate their results in comparison to existing software options. Additionally, we demonstrate how REGEM and METAGEM can be applied to improve GEI studies related to anthropometric traits in the UK Biobank and diabetes in the ProDiGY resource.

REGEM is a powerful tool that exploits the GEM methodology to enable rapid estimation of genome-wide summary statistics for any re-partition of a set of exposures and interaction covariates. One potential application of REGEM is in sensitivity analyses, a common epidemiological tool used to assess genetic confounding. In our analysis, we demonstrate that proper adjustment for interaction covariates can significantly reduce highly inflated summary statistics and increase the discovery of genetic loci. Such discoveries could have been missed due to the computational expense of repeated genome-wide calculations on individual-level data. While recent algorithms have enabled multi-threading capabilities,^2, 19^ high-performance computing, and cloud environments enable parallel genome-wide analysis, the pre-processing time required to set up these environments may add additional computational time and financial cost to individual-level genome-wide analysis. In our REGEM benchmark study, we show that by avoiding repeated computation on individual-level data, a genome-wide re-analysis can be completed within minutes, requiring minimal computation resources while still producing valid summary statistic results. REGEM is lightweight and can be run on local machines, greatly reducing runtime and cost compared to an equivalent individual-level data analysis.

Additionally, REGEM can also serve as a valuable pre-processing tool to harmonize summary statistics results from multiple GEI studies for downstream meta-analysis. This is particularly valuable in situations where different studies may test different combinations of exposure and interaction covariates. For instance, one study may jointly test G x sex and G x BMI, while another may only test G x sex. By applying REGEM to the first study, summary statistics from a model testing only G x sex can be obtained without having to re-analyze individual-level genotypes in that study. The resulting summary statistics from both studies can then be combined for meta-analysis without sharing individual-level data. Traditionally, harmonizing data from multiple GEI studies has been challenging due to lack of data sharing, privacy protection issues and logistics in data transportation and storage of individual-level data.^20^ Summary statistics-based algorithms help bypass such restrictions to facilitate collaborative research, and REGEM helps extend this family of tools to the GEI space.

Various GEI software programs can fit models with multiple interaction terms.^2, 19, 21^ However, limited statistical power remains a challenge, requiring larger study cohorts, especially in underrepresented populations.^22^ By enabling more flexible summary statistic-based meta-analysis, METAGEM provides an alternative strategy towards increasing overall sample size and statistical power for such analyses. For a single exposure meta-analysis without gene-by-covariate interactions, existing software options, such as the popular METAL program, are adequate. However, a nuanced set of considerations are required to determine whether it is appropriate to include additional terms in meta-analysis, whether related to additional exposure terms,^10^ gene-by-covariate interactions,^12^ or genetic main effects.^22^ For multiple interaction meta-analysis, METAGEM demonstrated efficient CPU time, though large memory space is required for larger numbers of interaction terms and unique variants across studies.

By facilitating more comprehensive, genome-wide analyses and meta-analyses involving interactions using only summary statistics, REGEM and METAGEM enable researchers to maximize the value of genome-wide interaction studies while minimizing computational time. A few limitations should be noted. Firstly, the GEM model corrects for standard covariates by removing them from the genotype and interaction matrices in a single projection step. While this approach improves computational performance of the primary GWIS considerably, it also takes away the possibility of modifying covariate main effect adjustments in subsequent re-analysis. Any such modification (e.g., seeking an interaction effect while completely removing a covariate main effect from the statistical model) would require a new analysis using individual-level data. Additionally, while REGEM has been shown to produce results that are consistent with those of GEM, improper GEI analysis using GEM, particularly in the case of rare variants, can lead to spurious summary statistics results, and may invalidate re-analysis results. Therefore, researchers must ensure valid summary statistics (for example, well-controlled genomic inflation) are generated from GEI methods before performing a re-analysis. In this vein, it is also important that study-specific interaction terms to be meta-analyzed have equivalent interpretations; for example, METAGEM cannot conduct valid meta-analysis when there are discrepant study-specific variable coding choices in terms of exposure (and covariate) centering.

In summary, we have introduced REGEM and METAGEM for further complex downstream analysis of GEI studies. REGEM and METAGEM, along with our GEM tool for genome-wide interaction analysis and corresponding workflows for reproducible and scalable deployment in cloud computing environments, are publicly available at (https://github.com/large-scale-gxe-methods). The suite of tools, including GEM, REGEM and METAGEM, provides key software infrastructure for maximizing the utility of summary statistics from diverse and complex GEI studies.

## Data Availability

The individual-level data that support the findings of this study are available upon application to the UK Biobank (https://www.ukbiobank.ac.uk/register-apply/).

## Acknowledgements

This research was conducted using the UK Biobank Resource under Application Numbers 27892 and 42646. This work was supported by NIH grant R01 HL145025. KEW was supported by NIH grant K01 DK133637. ProDiGY acknowledgements and funding sources are included in the Supplemental Material.

## Author contributions

D.T.P. and H.C. developed the METAGEM and REGEM algorithms. D.T.P., H.C., and C.P. implemented the METAGEM and REGEM software programs. D.T.P. and K.E.W. implemented software programs as cloud workflows. D.T.P. and H.C. designed the benchmark simulation study and carried out the analyses. K.E.W., L.C., and A.K.M. carried out the real-data analyses. S.S., E.I., M.E.V., F.B., S.C., R.G.-K., J.D., C.P., and S.M.M. provided guidance and input related to analysis of the ProDiGY dataset. K.E.W., D.T.P., H.C., and A.K.M. wrote the manuscript. All authors critically read the manuscript.

## Web resources

GEM, https://github.com/large-scale-gxe-methods/GEM

GEM Workflow, https://github.com/large-scale-gxe-methods/gem-workflow

METAGEM, https://github.com/large-scale-gxe-methods/METAGEM

METAGEM Workflow, https://github.com/large-scale-gxe-methods/metagem-workflow

REGEM, https://github.com/large-scale-gxe-methods/REGEM

REGEM Workflow, https://github.com/large-scale-gxe-methods/regem-workflow

## Data and code availability

METAGEM and REGEM are both open source projects freely available at https://github.com/large-scale-gxe-methods/METAGEM and https://github.com/large-scale-gxe-methods/REGEM. Workflows for both programs are also available at https://github.com/large-scale-gxe-methods/metagem-workflow and https://github.com/large-scale-gxe-methods/regem-workflow.

## References

1. Werme, J., van der Sluis, S., Posthuma, D., and de Leeuw, C.A. (2021). Genome-wide gene-environment interactions in neuroticism: an exploratory study across 25 environments. Transl. Psychiatry 11, 180. 10.1038/s41398-021-01288-9.

2. Westerman, K.E., Pham, D.T., Hong, L., Chen, Y., Sevilla-González, M., Sung, Y.J., Sun, Y.V., Morrison, A.C., Chen, H., and Manning, A.K. (2021). GEM: scalable and flexible gene-environment interaction analysis in millions of samples. Bioinformatics 37, 3514–3520. 10.1093/bioinformatics/btab223.

3. Bi, W., Zhao, Z., Dey, R., Fritsche, L.G., Mukherjee, B., and Lee, S. (2019). A Fast and Accurate Method for Genome-wide Scale Phenome-wide G × E Analysis and Its Application to UK Biobank. Am. J. Hum. Genet. 105, 1182–1192. 10.1016/j.ajhg.2019.10.008.

4. Gauderman, W.J., Zhang, P., Morrison, J.L., and Lewinger, J.P. (2013). Finding novel genes by testing G × E interactions in a genome-wide association study. Genet. Epidemiol. 37, 603–613. 10.1002/gepi.21748.

5. Kerin, M., and Marchini, J. (2020). Inferring Gene-by-Environment Interactions with a Bayesian Whole-Genome Regression Model. Am. J. Hum. Genet. 107, 698–713. 10.1016/j.ajhg.2020.08.009.

6. Mbatchou, J., Barnard, L., Backman, J., Marcketta, A., Kosmicki, J.A., Ziyatdinov, A., Benner, C., O’Dushlaine, C., Barber, M., Boutkov, B., et al. (2021). Computationally efficient whole-genome regression for quantitative and binary traits. Nat. Genet. 53, 1097– 1103. 10.1038/s41588-021-00870-7.

7. Zhong, W., Chhibber, A., Luo, L., Mehrotra, D.V., and Shen, J. (2023). A fast and powerful linear mixed model approach for genotype-environment interaction tests in large-scale GWAS. Brief. Bioinform. 24. 10.1093/bib/bbac547.

8. Shin, J., and Lee, S.H. (2021). GxEsum: a novel approach to estimate the phenotypic variance explained by genome-wide GxE interaction based on GWAS summary statistics for biobank-scale data. Genome Biol. 22, 183. 10.1186/s13059-021-02403-1.

9. Westerman, K., Liu, Q., Liu, S., Parnell, L.D., Sebastiani, P., Jacques, P., DeMeo, D.L., and Ordovás, J.M. (2020). A gene-diet interaction-based score predicts response to dietary fat in the Women’s Health Initiative. Am. J. Clin. Nutr. 111, 893–902. 10.1093/ajcn/nqaa037.

10. Kim, J., Ziyatdinov, A., Laville, V., Hu, F.B., Rimm, E., Kraft, P., and Aschard, H. (2019). Joint Analysis of Multiple Interaction Parameters in Genetic Association Studies. Genetics 211, 483–494. 10.1534/genetics.118.301394.

11. Moore, R., Casale, F.P., Jan Bonder, M., Horta, D., BIOS Consortium, Franke, L., Barroso, I., and Stegle, O. (2019). A linear mixed-model approach to study multivariate gene-environment interactions. Nat. Genet. 51, 180–186. 10.1038/s41588-018-0271-0.

12. Keller, M.C. (2014). Gene × environment interaction studies have not properly controlled for potential confounders: the problem and the (simple) solution. Biol. Psychiatry 75, 18–24. 10.1016/j.biopsych.2013.09.006.

13. Pan-UKB team. https://pan.ukbb.broadinstitute.org. 2020.

14. Willer, C.J., Li, Y., and Abecasis, G.R. (2010). METAL: fast and efficient meta-analysis of genomewide association scans. Bioinformatics 26, 2190–2191. 10.1093/bioinformatics/btq340.

15. Manning, A.K., Hivert, M.-F., Scott, R.A., Grimsby, J.L., Bouatia-Naji, N., Chen, H., Rybin, D., Liu, C.-T., Bielak, L.F., Prokopenko, I., et al. (2012). A genome-wide approach accounting for body mass index identifies genetic variants influencing fasting glycemic traits and insulin resistance. Nat. Genet. 44, 659–669. 10.1038/ng.2274.

16. TODAY Study Group, Zeitler, P., Epstein, L., Grey, M., Hirst, K., Kaufman, F., Tamborlane, W., and Wilfley, D. (2007). Treatment options for type 2 diabetes in adolescents and youth: a study of the comparative efficacy of metformin alone or in combination with rosiglitazone or lifestyle intervention in adolescents with type 2 diabetes. Pediatr. Diabetes 8, 74–87. 10.1111/j.1399-5448.2007.00237.x.

17. SEARCH Study Group (2004). SEARCH for Diabetes in Youth: a multicenter study of the prevalence, incidence and classification of diabetes mellitus in youth. Control. Clin. Trials 25, 458–471. 10.1016/j.cct.2004.08.002.

18. Srinivasan, S., Chen, L., Todd, J., Divers, J., Gidding, S., Chernausek, S., Gubitosi-Klug, R.A., Kelsey, M.M., Shah, R., Black, M.H., et al. (2021). The First Genome-Wide Association Study for Type 2 Diabetes in Youth: The Progress in Diabetes Genetics in Youth (ProDiGY) Consortium. Diabetes 70, 996–1005. 10.2337/db20-0443.

19. Chang, C.C., Chow, C.C., Tellier, L.C., Vattikuti, S., Purcell, S.M., and Lee, J.J. (2015). Second-generation PLINK: rising to the challenge of larger and richer datasets. Gigascience 4, 7. 10.1186/s13742-015-0047-8.

20. Reales, G., and Wallace, C. (2023). Sharing GWAS summary statistics results in more citations. Commun Biol 6, 116. 10.1038/s42003-023-04497-8.

21. Lin, D.-Y., Tao, R., Kalsbeek, W.D., Zeng, D., Gonzalez, F., 2nd, Fernández-Rhodes, L., Graff, M., Koch, G.G., North, K.E., and Heiss, G. (2014). Genetic association analysis under complex survey sampling: the Hispanic Community Health Study/Study of Latinos. Am. J. Hum. Genet. 95, 675–688. 10.1016/j.ajhg.2014.11.005.

22. Laville, V., Majarian, T., Sung, Y.J., Schwander, K., Feitosa, M.F., Chasman, D.I., Bentley, A.R., Rotimi, C.N., Cupples, L.A., de Vries, P.S., et al. (2022). Gene-lifestyle interactions in the genomics of human complex traits. Eur. J. Hum. Genet. 30, 730–739. 10.1038/s41431-022-01045-6.

