## Supplementary Material for "Summary statistics from large-scale gene-environment interaction studies for re-analysis and meta-analysis"

**Supplemental Material and Methods**

**ProDiGY Acknowledgements and Funding**

The TODAY and SEARCH for Diabetes in Youth studies are indebted to the many youth and families and the health care providers, whose participation made this study possible.

S.S. is supported by National Institute of Diabetes and Digestive and Kidney Diseases (NIDDK) grant K23DK120932. SEARCH 3/4 was supported by the South Carolina Clinical & Translational Research Institute at the Medical University of South Carolina, National Institutes of Health (NIH)/National Center for Advancing Translational Sciences (NCATS) grants UL1 TR000062 and UL1 TR001450; Seattle Children’s Hospital and the University of Washington, NIH/NCATS grant UL1 TR00423; University of Colorado Pediatric Clinical and Translational Research Center, NIH/NCATS grant UL1 TR000154; the Barbara Davis Center for Childhood Diabetes at the University of Colorado at Denver, Diabetes Endocrinology Research Centers NIH grant P30 DK57516; the University of Cincinnati, NIH/NCATS grants UL1 TR000077 and UL1 TR001425; and the Children with Medical Handicaps Program managed by the Ohio Department of Health. TODAY and TODAY Genetics was completed with funding from NIDDK and the NIH Office of the Director through grants U01-DK61212, U01-DK61230, U01-DK61239, U01-DK61242, and U01-DK61254; the National Center for Research Resources General Clinical Research Centers Program grants M01-RR00036 (Washington University School of Medicine), M01-RR00043-45 (Children’s Hospital Los Angeles), M01-RR00069 (University of Colorado Denver), M01-RR00084 (Children’s Hospital of Pittsburgh), M01-RR01066 (Massachusetts General Hospital), M01-RR00125 (Yale University), and M01-RR14467 (University of Oklahoma Health Sciences Center); and the National Center for Research Resources Clinical and Translational Science Awards grants UL1-RR024134 (Children’s Hospital of Philadelphia), UL1-RR024139 (Yale University), UL1-RR024153 (Children’s Hospital of Pittsburgh), UL1-RR024989 (Case Western Reserve University), UL1-RR024992 (Washington University in St. Louis), UL1-RR025758 (Massachusetts General Hospital), and UL1-RR025780 (University of Colorado Denver). SEARCH for Diabetes in Youth (SEARCH 3) is funded by the Centers for Disease Control and Prevention (PA numbers 00097, DP-05-069, and DP-10-001) and supported by the NIDDK. The SEARCH for Diabetes in Youth Cohort Study (1UC4DK108173) (SEARCH 4) is funded by the NIH/NIDDK and supported by the Centers for Disease Control and Prevention. The Population Based Registry of Diabetes in Youth Study (1U18DP006131, U18DP006133, U18DP006134, U18DP006136, U18DP006138, and U18DP006139) is funded by the Centers for Disease Control and Prevention and supported by the NIH/NIDDK. The sites for SEARCH 1–4 and funding are as follows: Kaiser Permanente Southern California (U18DP006133, U48/CCU919219, U01 DP000246, and U18DP002714), University of Colorado Denver (U18DP006139, U48/CCU819241-3, U01 DP000247, and U18DP000247-06A1), Cincinnati Children’s Hospital Medical Center (U18DP006134, U48/CCU519239, U01 DP000248, and 1U18DP002709), University of North Carolina at Chapel Hill (U18DP006138, U48/CCU419249, U01DP000254, and U18DP002708), Seattle Children’s Hospital (U18DP006136, U58/CCU019235-4, U01 DP000244, and U18DP002710-01), and Wake Forest University School of Medicine U18DP006131, U48/CCU919219, U01 DP000250, and 200-2010-35171). Sequencing for T2D-GENES cohorts was funded by NIDDK grant U01DK085526 (Multiethnic Study of Type 2 Diabetes Genes) and National Human Genome Research Institute grant U54HG003067 (Large Scale Sequencing and Analysis of Genomes). Sequencing for ProDiGY cohorts was funded by NIDDK grant U01DK085526. Analysis was supported by NIDDK grant U01DK105554 (AMP T2D-GENES Data Coordination Center and Web Portal). The Mount Sinai Institute for Personalized Medicine Biobank Program is supported by The Andrea and Charles Bronfman Philanthropies. The research from the Korean cohort was supported by a grant of the Korea Health Technology R&D Project through the Korea Health Industry Development Institute, funded by the Ministry of Health and Welfare, Republic of Korea (grants HI14C0060 and HI15C1595). The Framingham Heart Study is funded by NIH contracts N01-HC-25195 and HSN268201500001I and NIH grants U01 DK085526 and R01 DK078616. Additional support for the Framingham Heart Study is provided by the National Heart, Lung, and Blood Institute (NHLBI) (N01‐HC‐25195 and grant R01 NS17950) and National Institute on Aging (AG08122 and AG033193). This research was conducted in part using data and resources from the Framingham Heart Study of the NHLBI of the NIH and Boston University School of Medicine. The analyses reflect intellectual input and resource development from the Framingham Heart Study investigators participating in the SNP Health Association Resource project. This work was partially supported by the NHLBI’s Framingham Heart Study (contract number N01-HC-25195) and its contract with Affymetrix, Inc. for genotyping services (contract number N02-HL-6-4278). This research was partially supported by NIDDK grant 1R01DK8925601. A portion of this research used the Linux Cluster for Genetic Analysis. The Mexico City Diabetes Study has been supported by the following grants: RO1 HL24799 from the 2NHLBI; Consejo Nacional de Ciencia y Tecnología 2092, M9303, F677-M9407, 251M, 2005-C01-14502, and SALUD 2010-2151165; 1004 Type 2 Diabetes in Youth: ProDiGY Consortium Diabetes Volume 70, April 2021 and Consejo Nacional de Ciencia y Tecnología (Fondo de Cooperación Internacional en Ciencia y Tecnología C0012-2014-01-247974). The Diabetes in Mexico Study was supported by Consejo Nacional de Ciencia y Tecnología grant S008-2014-1-233970 and by Instituto Carlos Slim de la Salud, A.C.

This study includes data provided by the Ohio Department of Health, which should not be considered an endorsement of this study or its conclusions. The content is solely the responsibility of the authors and does not necessarily represent the official views of the NIH.

| Model | Exposure(s) | Interaction Covariate(s) |
| --- | --- | --- |
| Original | G x Sex | G x BMI |
| M1 | G x Sex, G x BMI | - |
| M2 | G x BMI | G x Sex |
| M3 | G x Sex | - |
| M4 | G x BMI | - |

**Supplemental Table 1**. Original model and variations of the original model (M1-M4) for the GEM and REGEM re-analysis benchmark study. All models were additionally adjusted for standard covariates: sex, BMI, age, age^2^, and PC1 – PC5.

| **Marker**  **Name** | **rs#** | **Nearest**  **gene** | **Non Effect Allele** | **Effect**  **Allele** | **Robust**  **Marginal**  **β (SE)** | **Robust G**  **β (SE)** | **Robust**  **G-age**  **β (SE)** | **Robust**  **G-sex**  **β (SE)** | **Robust**  **Marginal**  ***p*** | **Robust Interaction**  ***p*** | **Robust Joint**  ***p*** |
| --- | --- | --- | --- | --- | --- | --- | --- | --- | --- | --- | --- |
| ***Youth cases vs. Youth controls*** | |  |  |  |  |  |  |  |  |  |  |
| chr1:13983344:G:A | rs138172416 | *KAZN* | g | a | 0.65 (0.031) | 0.76 (0.16) | 0.19 (0.068) | 0.099 (0.38) | 0.00021 | 0.017 | **3.03×10^-8^** |
| chr5:127752693:C:T | rs567720919 | *CCDC192* | c | t | 0.82 (0.027) | 0.86 (0.16) | 0.12  (0.059) | -0.16 (0.34) | 4.64×10^-7^ | 0.099 | **1.72×10^-8^** |
| chr10:112998590:C:T | rs7903146 | *TCF7L2* | c | t | 0.47 (0.0052) | 0.48 (0.073) | 0.031 (0.026) | -0.017 (0.15) | **9.90×10^-11^** | 0.46 | **1.30×10^-9^** |
| chr12:4410686:C:T | rs114578532 | *FGF6* | c | t | 0.92 (0.027) | 0.88 (0.16) | -0.025 (0.050) | -0.26 (0.34) | **2.18×10^-8^** | 0.68 | 7.25×10^-7^ |
| chr12:17247195:A:AT | rs200936549 | *LMO3* | a | at | 0.58 (0.033) | 0.71 (0.17) | 0.018 (0.061) | 1.46 (0.42) | 0.0015 | 0.0019 | **1.09×10^-10^** |
| chr13:58252607:T:C | rs9537978 | *LINC00374* | t | c | 0.36 (0.0088) | 0.44 (0.092) | 0.11 (0.034) | 0.58 (0.19) | 0.00013 | 2.90×10^-5^ | **2.81×10^-9^** |
| chr15:53092645:A:G | rs115172264 | *RP11-209E8.1* | a | g | 0.36 (0.039) | 0.53 (0.19) | -0.019 (0.067) | 1.71 (0.43) | 0.072 | 0.00031 | **4.89×10^-8^** |
| chr15:77872306:G:C | rs114143681 | *GOLGA6FP* | g | c | 0.72 (0.037) | 0.85 (0.17) | -0.0034 (0.064) | 1.00 (0.41) | 0.00018 | 0.053 | **1.36×10^-8^** |
| chr18:60492770:TGTGTGTGTGC:T | rs1479869222 | *MC4R* | tgtgtgtgtgc | t | 0.51 (0.036) | 0.63 (0.17) | 0.0016 (0.050) | 1.41 (0.44) | 0.0070 | 0.0059 | **9.22×10^-9^** |

**Supplemental Table 2:** Top signals from meta-analyses in the youth (youth cases vs. youth controls) subset of the ProDiGY cohort.

| **Marker**  **Name** | **rs#** | **Nearest**  **gene** | **Non Effect Allele** | **Effect**  **Allele** | **Robust**  **Marginal**  **β (SE)** | **Robust G**  **β (SE)** | **Robust**  **G-age**  **β (SE)** | **Robust**  **G-sex**  **β (SE)** | **Robust**  **Marginal**  ***p*** | **Robust Interaction**  ***p*** | **Robust Joint**  ***p*** |
| --- | --- | --- | --- | --- | --- | --- | --- | --- | --- | --- | --- |
| **Adult cases vs. Adult controls** | |  |  |  |  |  |  |  |  |  |  |
| chr3:156287496:G:A | rs62287662 | *KCNAB1* | g | a | -0.32 (0.016) | -0.35 (0.13) | 0.059 (0.010) | 0.036 (0.25) | 0.010 | 6.27×10^-8^ | 1.79×10^-8^ |
| chr10:112995025:T:C | rs35198068 | *TCF7L2* | t | c | 0.26 (0.0012) | 0.26 (0.034) | -0.0014 (0.0029) | -0.039 (0.068) | 2.01×10^-14^ | 0.89 | 1.23×10^-12^ |
| chr11:2818521:C:T | rs2237892 | *KCNQ1* | c | t | -0.28 (0.0016) | -0.28 (0.040) | -0.0034 (0.0035) | -0.024 (0.080) | 4.73×10^-12^ | 0.60 | 1.03×10^-10^ |

**Supplemental Table 3:** Top signals of meta-analyses in the adult (adult cases vs. adult controls) subset of the ProDiGY cohort.

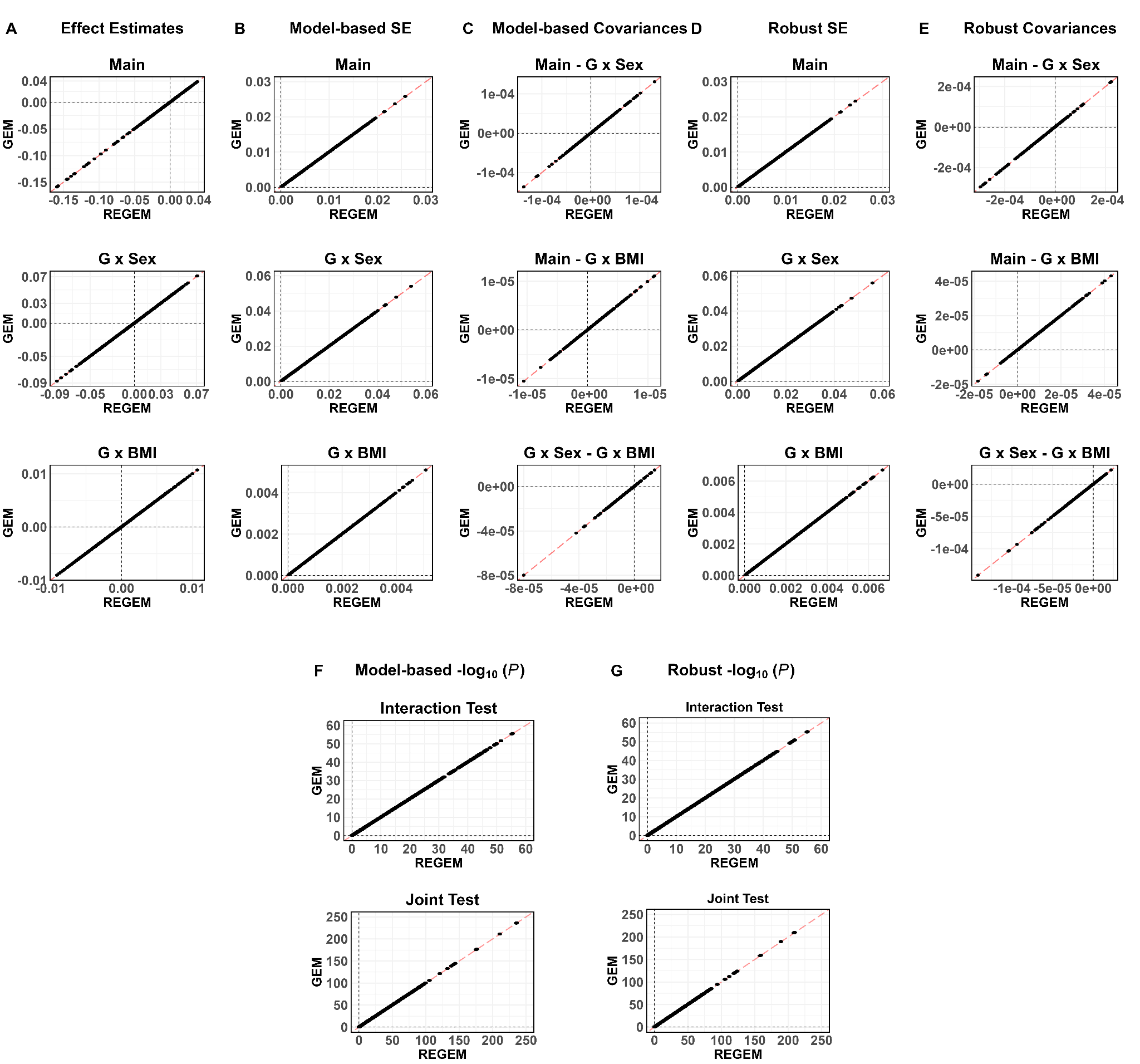

**Supplemental Figure 1:** Genome-wide summary statistics comparisons between GEM and REGEM from the joint G x Sex and G x BMI GEI test (M1). In each plot, the results of REGEM are on the x-axis and results for GEM are on the y-axis. The diagonal dark red dashed line is the identity line. (A-E), the columns represent the effect estimates, model-based standard errors, model-based covariances, robust standard errors, and robust covariances comparisons for the main, G x Sex, and G x BMI terms in the first, second, and third rows, respectively. In (F), the rows represent the model-based interaction and joint tests *P*-value comparisons in the -log_10_ scale. In (G), the rows represent the robust interaction and joint tests *P*-value comparisons in the -log_10_ scale.

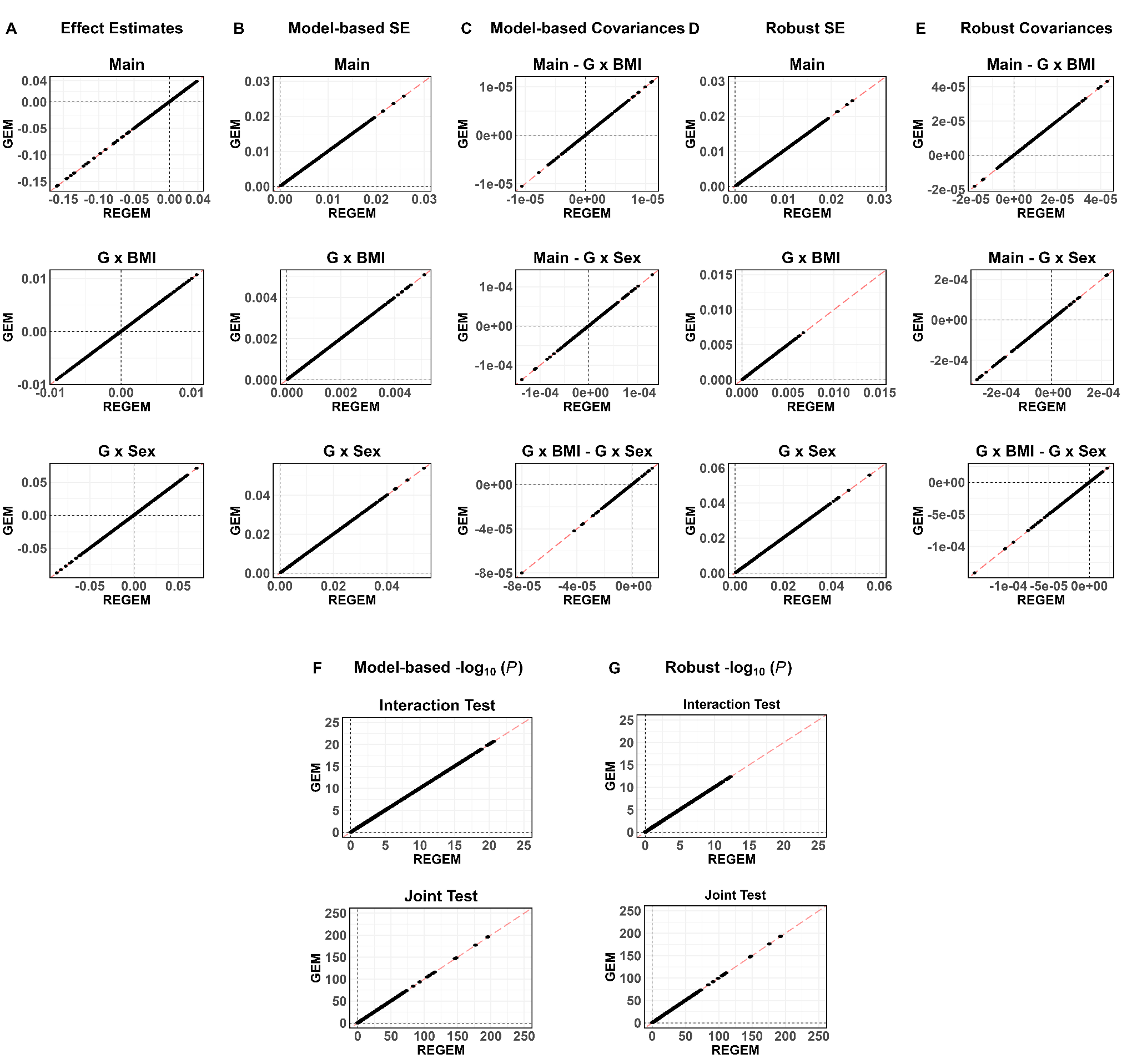

**Supplemental Figure 2:** Genome-wide summary statistics comparisons between GEM and REGEM from testing G x BMI while adjusting for G x Sex (M2). In each plot, the results of REGEM are on the x-axis and results for GEM are on the y-axis. The diagonal dark red dashed line is the identity line. (A-E), the columns represent the effect estimates, model-based standard errors, model-based covariances, robust standard errors, and robust covariances comparisons for the main, G x BMI, and G x Sex terms in the first, second, and third rows, respectively. In (F), the rows represent the model-based interaction and joint tests *P*-value comparisons in the -log_10_ scale. In (G), the rows represent the robust interaction and joint tests *P*-value comparisons in the -log_10_ scale.

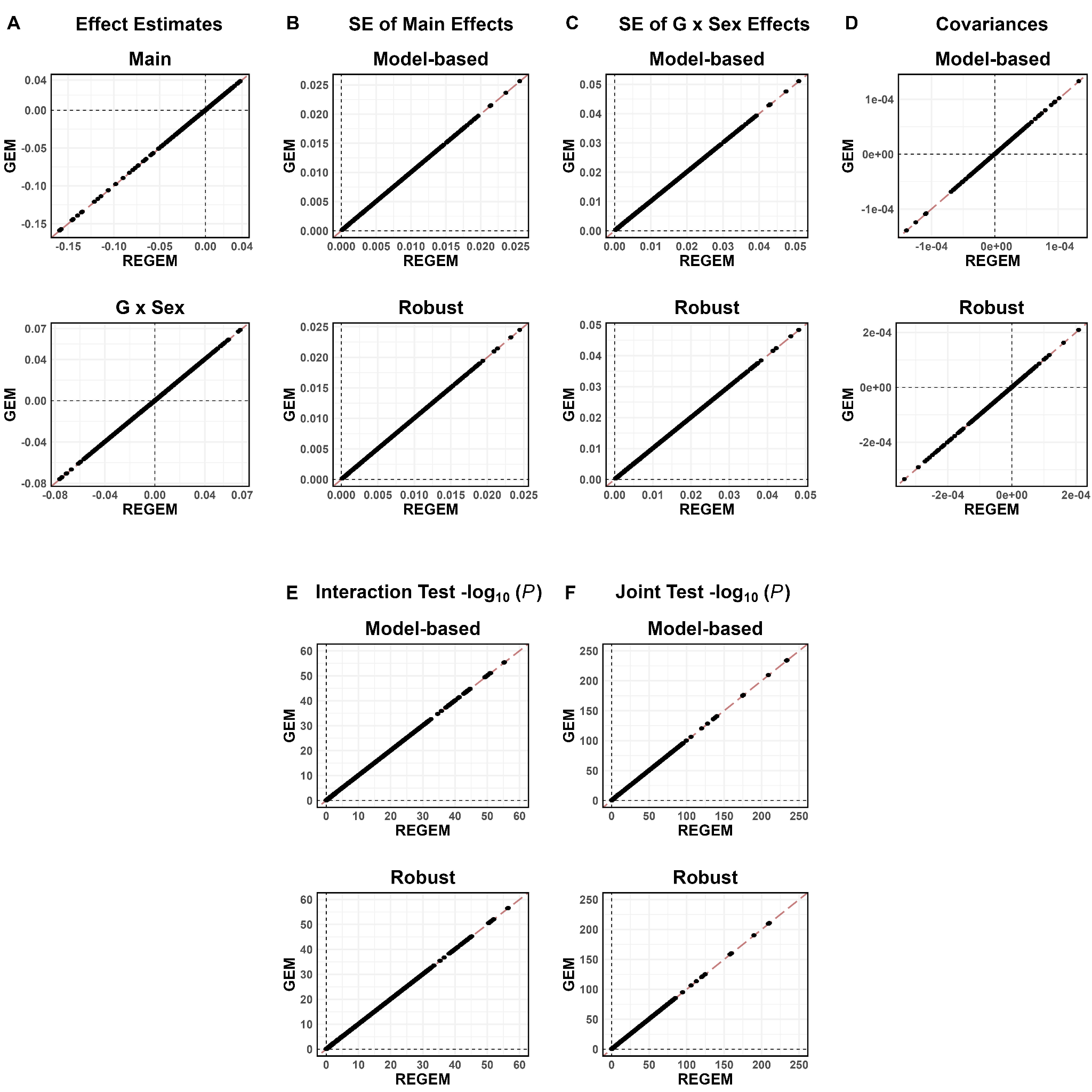

**Supplemental Figure 3:** Genome-wide summary statistics comparisons between GEM and REGEM from testing G x Sex (M3). In each plot, the results of REGEM are on the x-axis and results for GEM are on the y-axis. The diagonal dark red dashed line is the identity line. (A) the main and G x Sex effect estimates comparisons on the first and second row, respectively. In each column of B-F, the first and second rows are the model-based and robust summary statistics comparisons. (B) standard errors of the main effect estimates. (C) standard errors of the G x Sex effect estimates. (D) covariance between the main and G x Sex effect estimates. (E) comparison between the interaction test *P*-values in the -log_10_ scale. (F) comparison between the joint test *P*-values in the -log_10_ scale.

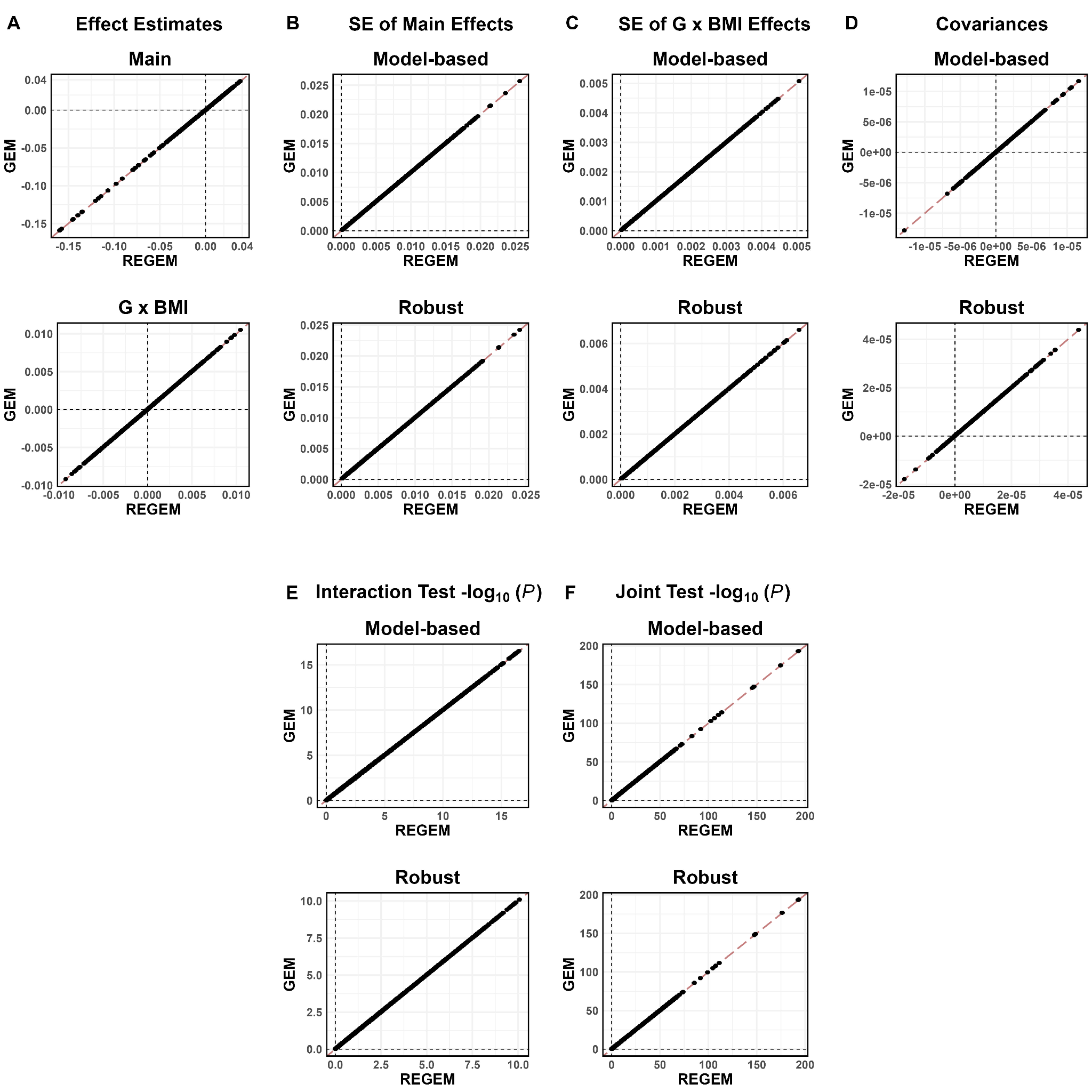

**Supplemental Figure 4:** Genome-wide summary statistics comparisons between GEM and REGEM from testing G x BMI (M3). In each plot, the results of REGEM are on the x-axis and results for GEM are on the y-axis. The diagonal dark red dashed line is the identity line. (A) the main and G x BMI effect estimates comparisons on the first and second row, respectively. In each column of B-F, the first and second rows are the model-based and robust summary statistics comparisons. (B) standard errors of the main effect estimates. (C) standard errors of the G x BMI effect estimates. (D) covariance between the main and G x BMI effect estimates. (E) comparison between the interaction test *P*-values in the -log_10_ scale. (F) comparison between the joint test *P*-values in the -log_10_ scale.

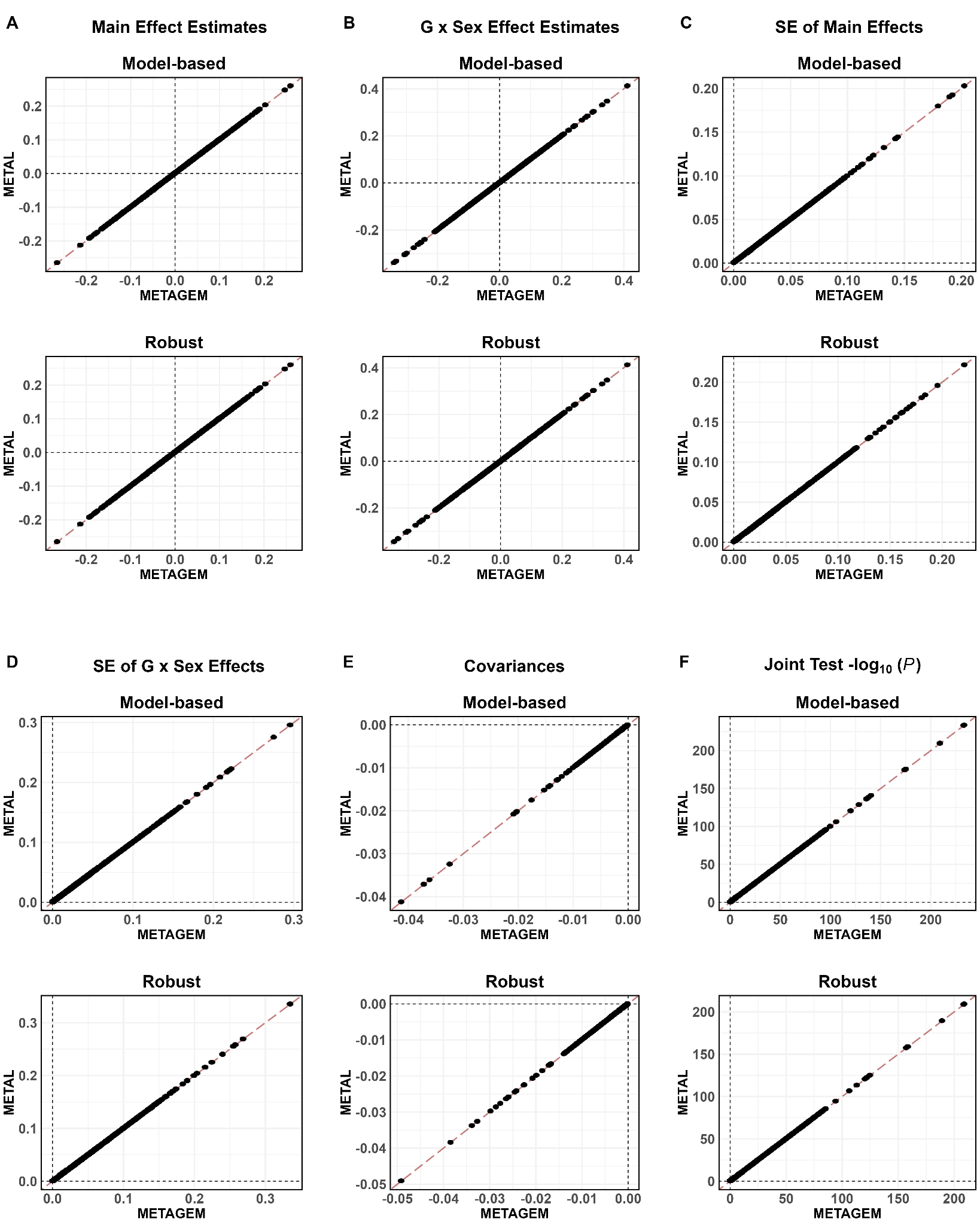

**Supplemental Figure 5:** Genome-wide meta-analysis comparisons between METAGEM and METAL summary statistics results from the sex interaction test. In each column A-F, the first and second rows are the model-based and robust result comparisons, respectively. In each plot, results for METAGEM are shown on the x-axis and results for METAL are on the y-axis. The diagonal dark red dashed line is the identity line. (A) main effect estimates. (B) G x Sex effect estimates. (C) standard errors of the main effect estimates. (D) standard errors of the G x Sex effect estimates. (E) covariances between the main and G x Sex effect estimates. (F) The meta-analysis joint test *P*-values in the -log_10_ scale.

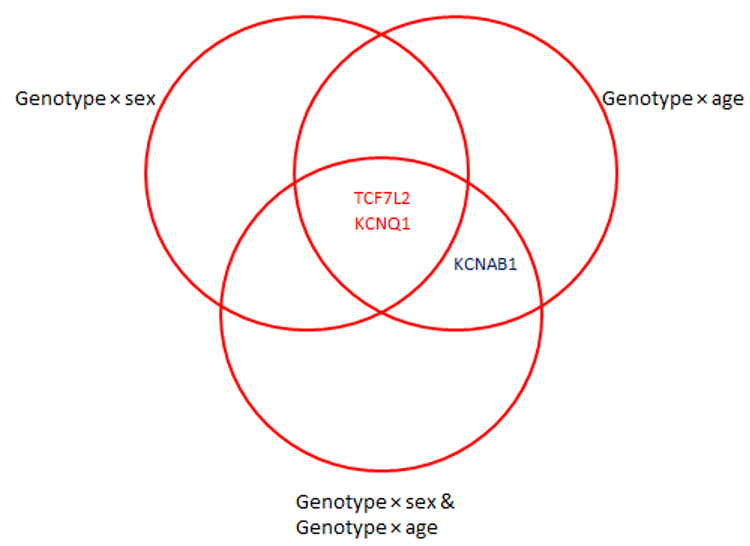

**Supplemental Figure 6:** Results from multi-exposure GWIS for incident T2D in the ProDiGY adult cohort. Venn diagram displays overlap between loci discovered at genome-wide significance using the joint test of genetic and interaction effects (*p_joint_* = 5×10^-8^), from each of: sex-only, age-only, and multi-exposure (sex and age) analyses. Variants are labeled according to the closest gene, and colors correspond to the test(s) in which significance was achieved: joint genetic effect (dark blue), or both joint and marginal genetic effects (red).
